# Bereavement care for ethnic minority communities: A systematic review of access to, models of, outcomes from, and satisfaction with, service provision

**DOI:** 10.1101/2021.02.13.21251679

**Authors:** Catriona Rachel Mayland, Richard A. Powell, Gemma Clarke, Bassey Ebenso, Matthew J Allsop

**Affiliations:** Department of Oncology and Metabolism, University of Sheffield, Sheffield, UK; Sheffield Teaching Hospitals NHS Foundation Trust, Sheffield, UK; Palliative Care Department, University of Liverpool, Liverpool, UK; School of Public Health, Faculty of Medicine, Imperial College London, UK; Academic Unit of Palliative Care, Leeds Institute of Health Sciences, University of Leeds, UK; Nuffield Centre for International Health and Development, Leeds Institute of Health Sciences, University of Leeds, UK

**Keywords:** Bereavement, grief, minority groups, ethnic groups, systematic review

## Abstract

**Objectives:** To review and synthesize the existing evidence on bereavement care, within the United Kingdom (UK), for ethnic minority communities in terms of barriers and facilitators to access; models of care; outcomes from, and satisfaction with, service provision.

**Design:** A systematic review adopting a framework synthesis approach was conducted. An electronic search of the literature was undertaken in MEDLINE, Embase, PsycINFO, Social Work Abstract and CINAHL via EBSCO, Global Health, Cochrane library, the Trip database and ProQuest between 2000 and 2020. Search terms included bereavement care, ethnic minority populations and the UK setting.

**Results:** From 3,185 initial records, following screening for eligibility, and full-text review of 164 articles, seven studies were identified. There was no research literature outlining the role of family, friends and existing networks; and a real absence of evidence about outcomes and levels of satisfaction for those from an ethnic minority background who receive bereavement care. From the limited literature, the overarching theme for barriers to bereavement care was ‘unfamiliarity and irregularities’. Four identified subthemes were ‘lack of awareness’; ‘variability in support’; ‘type and format of support’; and ‘culturally specific beliefs’. The overarching theme for facilitators for bereavement care was ‘accessibility’ with the two subthemes being ‘readily available information’ and ‘inclusive approaches’. Three studies reported on examples of different models of care provision.

**Conclusions:** This review reveals a stark lack of evidence about bereavement care for ethnic minority populations. In particular, understanding more about the role of family, friends and existing support systems, alongside outcomes and satisfaction will begin to develop the evidence base underpinning current provision. Direct user-representation through proactive engagement and co-design approaches may begin to determine the most appropriate models and format of bereavement care for ethnic minority communities to inform service design and delivery.

## Introduction

By 2^nd^ January 2021, 51,437 individuals had died from COVID-19 within the United Kingdom (UK).(1) A growing concern is the disproportionate impact of the virus on people from ethnic minority communities. This includes impact within both the healthcare sector and in the general population, possibly from situational vulnerabilities (i.e. socio-economic disadvantage, manifesting in factors such as greater exposure to infection and higher prevalence of health vulnerability).(2) Among over 60,000 excess UK deaths occurring during the pandemic, the highest death rates are recorded among ethnic minority groups.(3) With each decedent potentially affecting at least five others(4), this reflects a substantial number affected by grief and bereavement.

Experiences of death and bereavement are likely to be significantly affected on both an individual and societal level during the pandemic influenced by factors including the nature of the death, existing family and social support networks and cultural context.(5) Compared with the causes of ‘typical’ deaths, COVID-19 has distressing symptoms, a rapid progression to the end of life in severe cases, distress related to patient and family isolation (in care settings, and in bereavement),(6) and enforced limitations on meaningful end-of-life rituals. This multiplicity of losses associated with pandemics impacts upon cultural norms, rituals, and usual social practices related to death and mourning, potentially increasing the risk of complicated grief.(7)

A recent mapping of ethnic minority mental health services in the UK(8) noted the need to ensure anyone bereaved by COVID-19 receive the appropriate support they need. It was recognised, however, that ethnic minority groups are less likely to access mainstream bereavement services(8). This may relate to access issues, as well as services not being culturally sensitive or designed in a way to meet the needs of specific communities.(9) One of the key recommendations from this recent report is the need to address gaps in research surrounding ethnicity, bereavement and loss in the UK.(8)

In order to identify gaps in research about the existing provision of bereavement support for ethnic minority groups, the public health model(10) provides a conceptual framework aligning interventions with need across three groups. The model proposes that for the majority of individuals, their own inner resources, family and friends, will support their distress and subsequent adjustments to their losses. Approximately a third need non-specialist, informal support. Only a small proportion are at risk of developing ‘complicated grief,’ characterised by intense grief, which can last for a longer period compared with social expectations or cause impairment in daily functioning(11). The differing components of bereavement support are defined into tiers according to the level of need (12) (Textbox 1):

### Textbox 1.

**Components of bereavement support (National Bereavement Alliance, NBA)**

- **Component 1:** most support provided by family, friends, existing networks; information about the experience of bereavement and sign posting to further support is offered on a universal basis (*approximately sixty percent*).
- **Component 2:** individual or group-based structured support sessions (faith groups, befriending groups) for those who are seeking that type of support or are at risk of developing more complex needs (*approximately thirty percent*).
- **Component 3:** specialist interventions provided by specialist counsellors, psychologists or mental health practitioners for those with complex needs, pre-existing mental health conditions or are at high risk of developing prolonged or complicated grief (*approximately ten percent*).

It is important to acknowledge potential limitations of this public health model from the outset, however, in terms of its applicability to ethnic minority communities. The model was developed by an Australian team of researchers, and initial pilot data involved bereaved carers predominately from an ‘Australian’ and ‘other English speaking’ background.(13) Terms such as ‘bereavement’ and ‘grief’ are quite specific to the English language and can be difficult to translate into languages which may not have specific words to link with emotions associated with loss or death.(14) A pragmatic approach, however, was taken to use this framework to guide initial exploration and mapping of research reporting bereavement support for ethnic minority populations in the UK.

This systematic literature review seeks to synthesize the existing evidence on bereavement care for ethnic minority populations through addressing the following research questions:

1. What are the barriers and facilitators to accessing bereavement care for affected people from ethnic minority populations?
2. What, if any, models of care provision exist to address specific ethnic minority population needs around bereavement?
3. What outcomes are reported for ethnic minority populations accessing bereavement services?
4. What are the levels of satisfaction with bereavement services reported by bereaved people from ethnic minority populations?

In order to clarify what is being described, the following definitions will be used:

- Bereavement care: all care provided to those bereaved (all three components of the National Bereavement Alliance (NBA) model) including support from family, friends and existing networks.
- Bereavement support: interventions provided at components one and two (provision of information, informal support, individual or group-based structured support).
- Bereavement counselling: support provided at component three (specialist interventions).

It is anticipated that findings will clarify the extent of the existing evidence base and potential gaps as well as provide guidance upon which future research, service design and planning, and resource allocation for UK bereavement services can be built.

## Methods

This review adopted a framework synthesis approach, enabling a structured approach to both organizing and interpreting data.(15) Prior to conducting the review, the research team developed a protocol to guide the review, outlining details of the inclusion criteria and databases to search. The review is reported according to the Preferred Reporting Items for Systematic Reviews and Meta-Analyses (PRISMA) guidelines.(16)

### Data Sources and Searches

Databases searched included MEDLINE, Embase, PsycINFO, Social Work Abstracts and CINAHL via EBSCO, Global Health, Cochrane library, the Trip database and ProQuest. This approach reflects a wide range of databases relevant to social science, medical, nursing and allied health professionals and includes grey literature. We conducted database searches on 19^th^ August 2020. The search strategy included terms for bereavement care, ethnic minority populations (including religions commonly practiced by ethnic minority groups in the UK), and terms to focus the review on the UK setting. An example of the search strategy used for MEDLINE is provided in Supplementary File 1.

### Study Selection

Studies were included if they reported on bereaved participants of any age (both adults and children) from UK ethnic minority populations or caregivers/family members bereft from death of a person from an ethnic minority group (population). The definition of ‘ethnic minority population’ is aligned with the UK Office for National Statistics, comprised of all ethnic groups other than White British, and is inclusive of minority White ethnic groups. Included studies covered any study design and published in peer-reviewed journals. Studies were excluded if they did not report data specific to ethnic minority populations or were not conducted with a focus on the UK setting. Studies were limited to those published in the last 20 years.

### Data Extraction

BE, GC and MJA independently screened the titles and abstracts of studies identified. Discrepancies in screening inclusion and exclusion were resolved through discussion. Data were extracted from each study on the following sample and methodological characteristics: study design, number of participants, summaries of age, gender and ethnicity of study participants, components of bereavement support described, and a summary of key findings.

### Quality Assessment

Due to the heterogeneity of study types, the Mixed Method Appraisal Tool (MMAT) was used for assessing the quality of both quantitative and qualitative studies(17). All studies were appraised using the MMAT checklist to rate the methodology and rigour by GC and MJA. Study quality was assessed to appraise included studies but did not inform any exclusion.

### Data Synthesis and Analysis

A framework synthesis(15) approach was adopted, incorporating five stages of familiarisation, framework selection, indexing, charting and mapping and interpretation. An initial framework reflecting key components relating to a person’s access and interaction with bereavement care across the three levels of the NBA tiers was developed using two sources: literature on known inequities in provision of palliative care (e.g. access, quality and experience of care)(18) alongside expertise across the team in health services and bereavement research. This included four components of: i) access; ii) models and approaches to delivery and provision of bereavement care; iii) outcomes of those accessing bereavement services; and iv) satisfaction with or appraisal of care received. Data from the results and discussion sections of included articles were independently coded by two authors (MJA, CM) and aligned with the four components of the framework. The framework was adapted and refined during the analysis of data, with documentation of data arising that augmented the definition and content of the four categories. Following alignment of the included studies and meetings between researchers to discuss interpretation and analysis of study findings, an initial conceptual framework was developed. The framework was used to inform engagement with three patient representatives from ethnic minority populations in Bradford and Sheffield and a range of key stakeholders from national advocacy and third sector organisations. This engagement sought to elicit feedback on the framework, guide appraisal of the analysis based on their own experiences and incorporate feedback on alignment of data informed by their expert knowledge.

## Results

From 3,185 initial records, 3,085 papers were screened for eligibility with full-text reviews of 164 articles reviewed, of which seven were included (Figure 1). These were published across a 20-year period, with two articles published since 2010. Most studies (n=5) adopted a descriptive observational approach, including a retrospective data analysis(19), questionnaire surveys(20, 21), and face-to-face interviews and focus group discussions(22). Analytic observational studies were adopted in two studies using comparative cross-sectional questionnaire face-to-face surveys(23, 24); these two articles report data from the same dataset.

**Figure 1:**
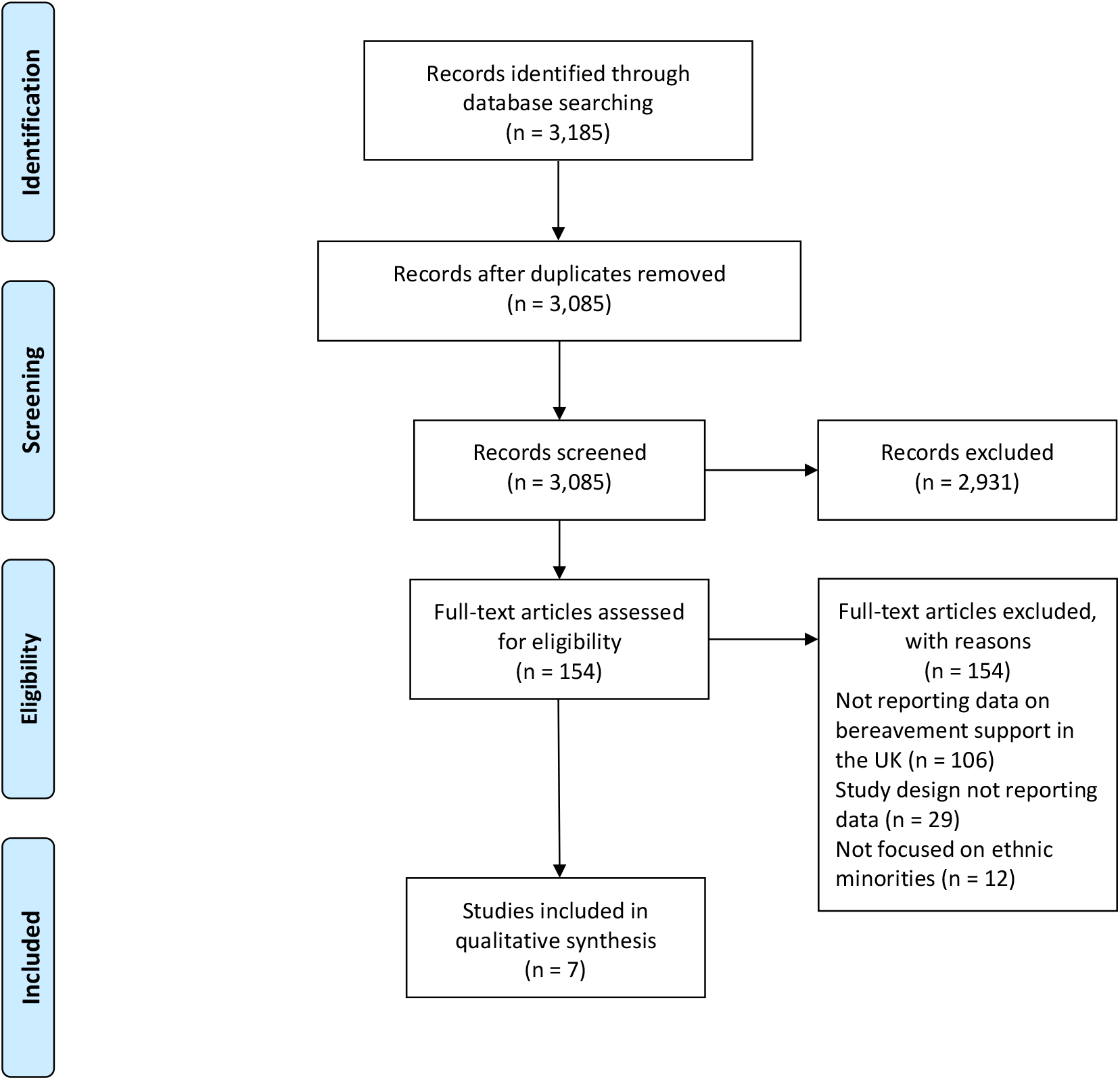
PRISMA flow chart outlining screening and inclusion of articles in the review.

Four of the studies were conducted in London with a focus on specific local authorities(20, 23-25). Bradford was the location of one study(19). The remaining two had a national focus with recruitment taking place across England(22), and a study surveying neonatal units across the UK(21).

Studies were focused across secondary and community care settings. Two secondary care studies focused on hospital-based palliative care teams and included one(19) and multiple(20) sites. A third secondary care study focused on hospital-based neonatal teams(21). Community-based studies included a focus on bereavement among family or close friends of first-generation black Caribbean and white populations(23, 24), exploration of bereavement in the context of Gypsy and Traveller communities(22), and accounts of bereaved carers of Bangladeshi patients in east London and their interaction with community-based palliative care(25).

### Quality appraisal

Across all studies, most criteria were met for the relevant MMAT checklists (Supplementary file 2). For qualitative studies(22, 25), 100% of quality criteria were reported. For quantitative descriptive studies(19-21), all applicable criteria were met for two studies(19, 21), it was not possible to determine the relevance of the sampling strategy, sample representativeness or risk of nonresponse bias for a component of a study assessing memorial service invitations(20). For quantitative non-randomised studies,(23, 24) both reporting data from the same dataset, one met all quality criteria (23). The second of the two quantitative non-randomised studies, which was a short focused report, did not contain information to determine whether confounders were accounted for in the design and analysis (24).

### Participant characteristics

Participants included caregivers, relatives and friends (n=185) and service providers (n=389) (Table 1). Where reported, age of caregivers was mostly under 54 years (80% of participants) for two studies reporting data on black Caribbean participants (23, 24) and a median age of 35 for bereaved Bangladeshi caregivers (25).

**Table 1:** Data on study population extracted from included studies.

| Citation | Population description | Participant description | Age | Gender | Race/ethnicity composition | How identified as ethnic minorities |
| --- | --- | --- | --- | --- | --- | --- |
| 18 | Patients from ethnic minorities referred to the specialist Bradford teaching hospitals palliative care team from 1 October 2001 to 30 September 2002 were identified using the hospital palliative care team database. | 47 patients | Not specified | 30 were male (64%) and 17 were female (36%) | African (n=1), Afro-Caribbean (n=5), Bangladeshi (n=3), Eastern European (n=8), Indian (n=6), Pakistani (n=23) | Review of medical records for demographic information (sex, age, disease, religion and country of origin), the patient's first language, whether they could speak English and whether an interpreter was advised on referral to the team. |
| 22 | Three materially and socially deprived London local authorities - Black Caribbean and White British relatives who registered a death | 100 family members or close friends | Black Caribbean (80% under 54 years) and White British (48% under 54 years) | 72% of Black Caribbean participants were female; 62% of White British participants were female | Black Caribbean (N= 50) White British (N= 50) | Country of origin registered on death certificate - validated at interview |
| 23 | Three materially and socially deprived London local authorities - Black Caribbean and White British relatives who registered a death | 100 family members or close friends | Black Caribbean (80% under 54 years) and White British (48% under 54 years) | 72% of Black Caribbean participants were female; 62% of White British participants were female | Black Caribbean (N= 50) White British (N= 50) | Country of origin registered on death certificate - validated at interview |
| 19 | Staff of bereavement teams based in hospital palliative care teams | 59 nursing staff | Not reported | Not Reported | Not reported | Not applicable |
| 20 | Bereavement teams in neonatal care centres in the UK | 330 staff in neonatal units (87 doctors; 133 nurses; 84 chaplains; 26 not stated) | Not reported | Not Reported | Not Reported | Not applicable |
| 21 | British Romany Gypsies and travellers | 20 bereaved relatives | Only age of relative at death is provided | All participants were female | Irish Travellers (n=13) and English Gypsies (n=7) | Via non-governmental organisations (NGOs) that provide advocacy and support to Gypsy/Traveller communities by promoting social inclusion and equality with mainstream society. |
| 24 | Bangladeshi caregivers | 18 bereaved caregivers | Median age of 35, ranging from 25 - 60 years old | 8 of 18 participants were female | Bangladeshi (n=18) | Not applicable |

Ethnic groups represented across the studies included Black Caribbean (N= 50), White British (N= 50), Pakistani (n=23), Bangladeshi (n=18), Irish Travellers (n=13), Eastern European (n=8), English Gypsies (n=7), Indian (n=6), Afro-Caribbean (n=5), Bangladeshi (n=3), and African (n=1).

### Service providers

Service provider participants included nursing staff based in the bereavement teams of hospital palliative care teams (n=59), alongside doctors (n=87), nurses (n=133), chaplains(n=84) and not specified (n=26) participants based in neonatal units across the UK. Age and ethnicity of service provider participants were not reported.

### Key findings from included studies

Table 2 outlines the data on study design, setting and key findings of included studies. The findings, as aligned to four components of the framework used to guide the analysis, are outlined below.

**Table 2:**
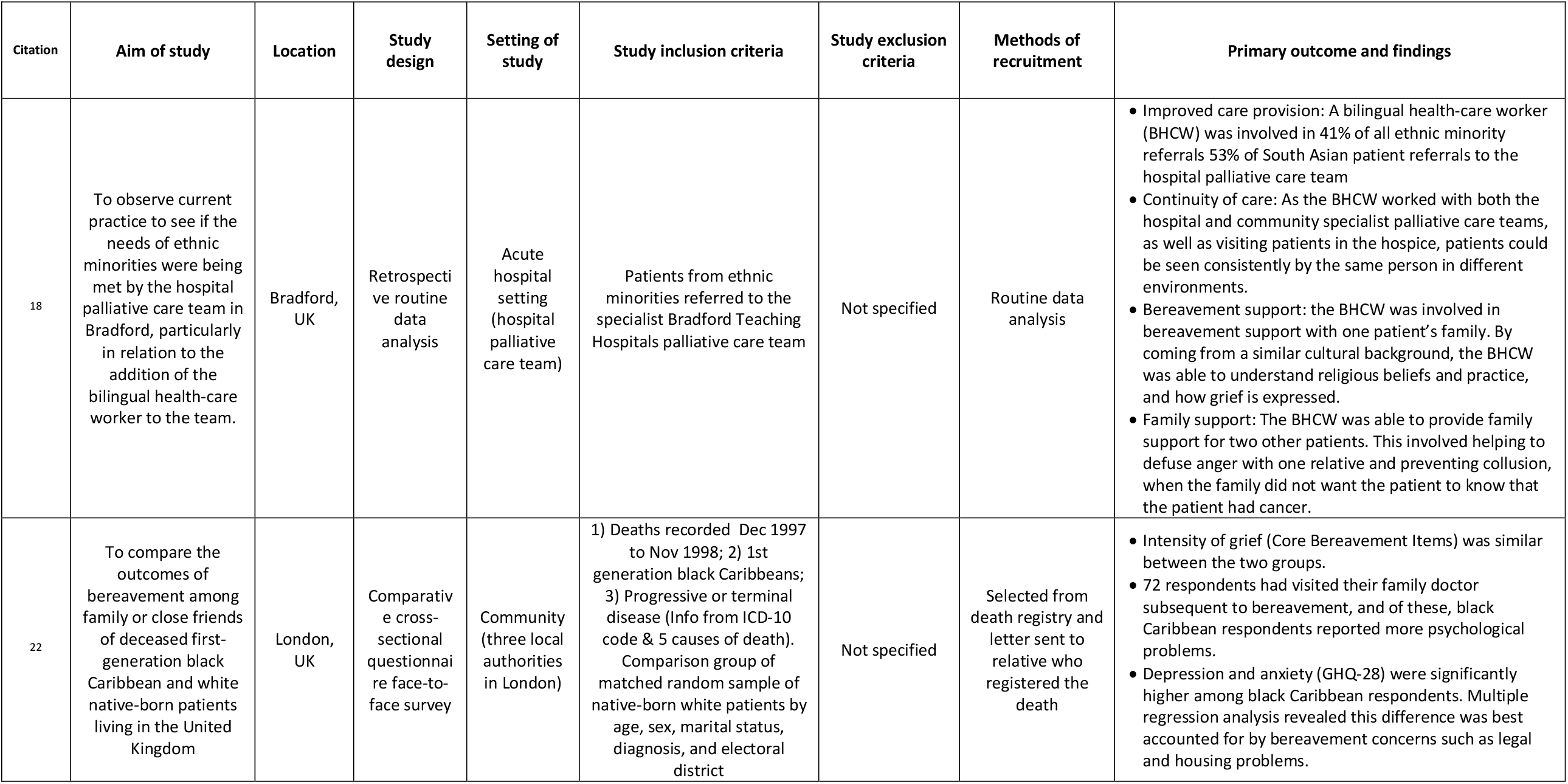
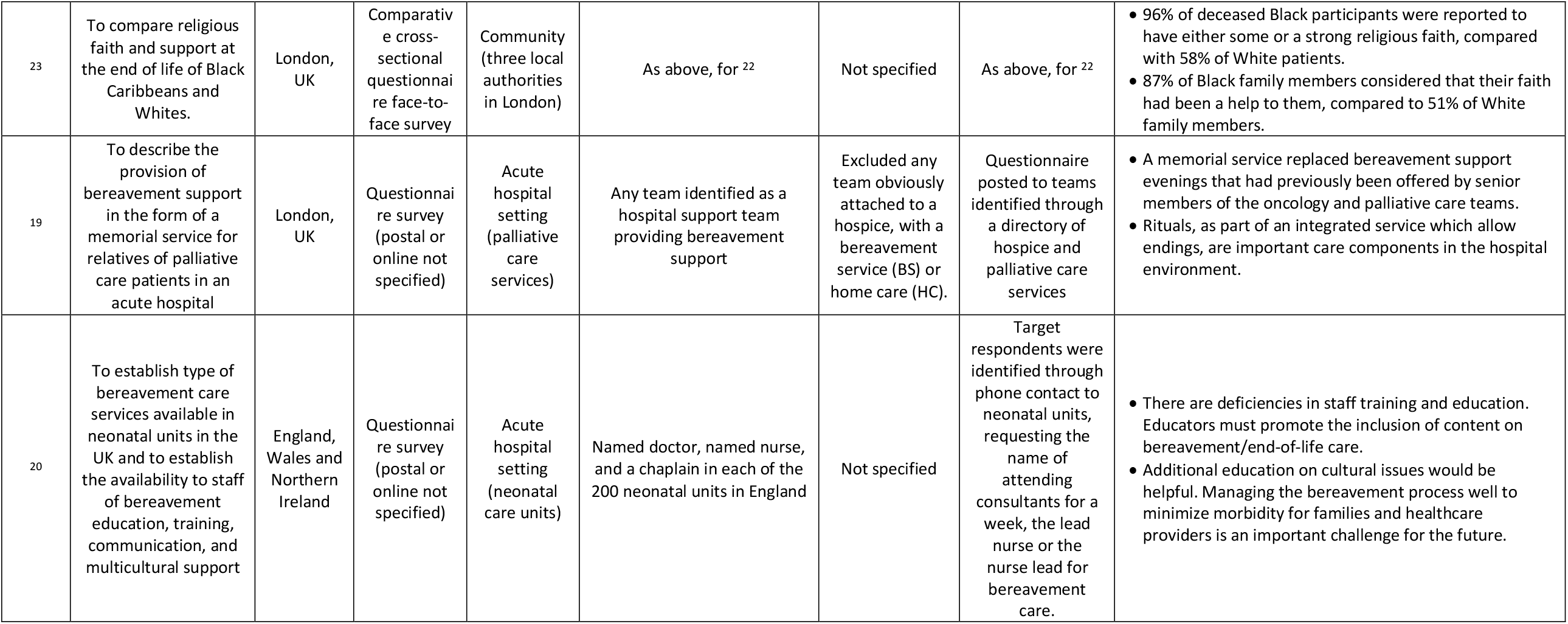
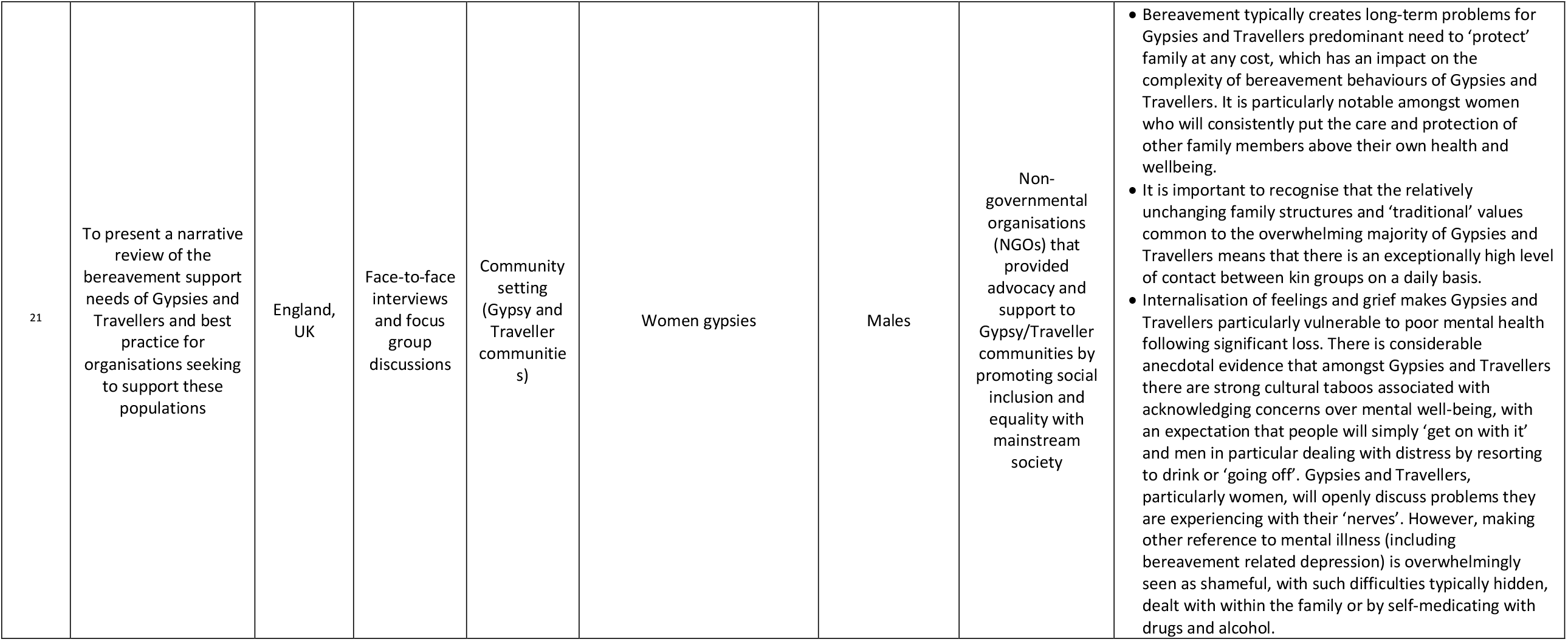
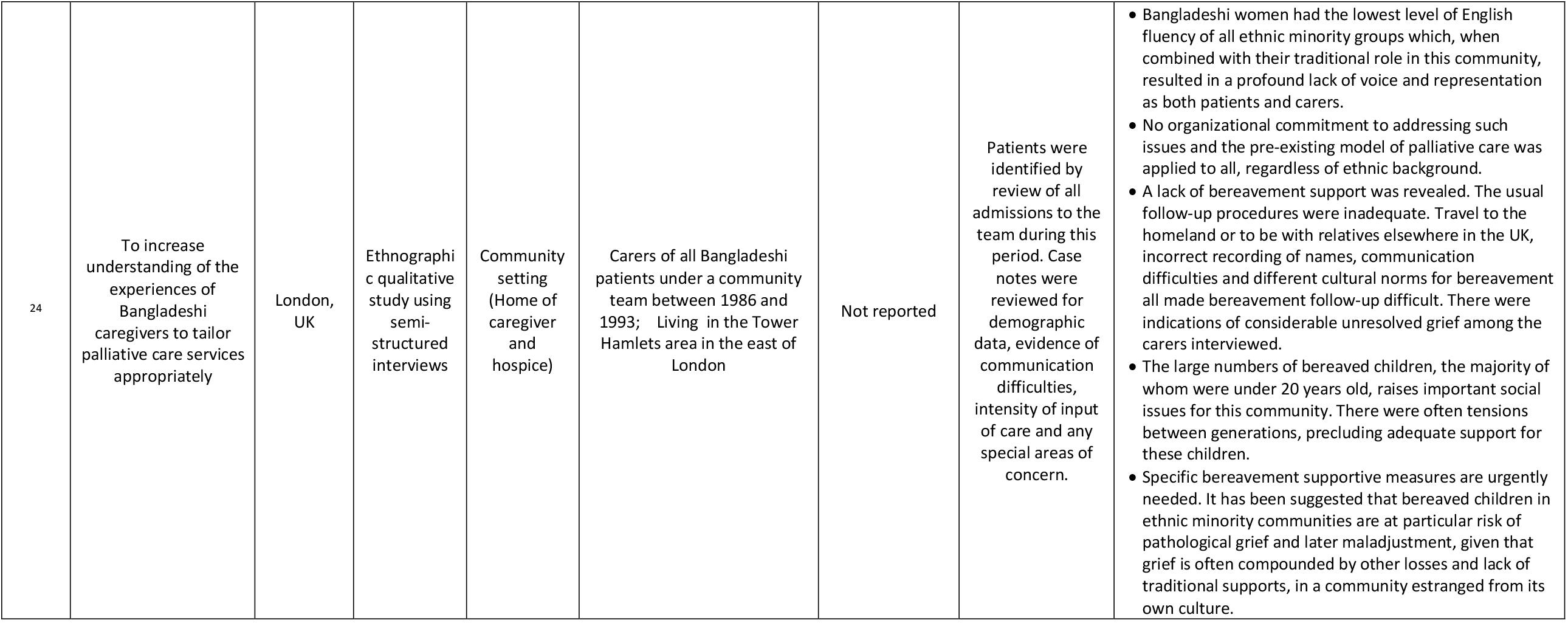
Data on study design, setting and key findings of included studies.

#### Access: barriers and facilitators to bereavement support services

There were six studies which included some aspect relating to barriers and/or facilitators to accessing bereavement care for people from ethnic minority populations.

#### Barriers

The overarching theme was ‘unfamiliarity and irregularities’ with four identified subthemes including ‘lack of awareness’; ‘variability in support’; ‘type and format of support’; and ‘culturally specific beliefs’.

The lack of awareness about bereavement care for ethnic minority communities could arise from factors relating to healthcare professionals. Limited training on bereavement care, perceptions about the training being inadequate (especially around cultural issues) and a dearth of readily available information packs about bereavement care were reported issues.(21) Additionally, medical practitioners’ tendency to prescribe medication, rather than provide information and offer psychological counselling, resulted in a lack of awareness about the wider support that could be offered for Romany Gypsy and Traveller communities.(22)

From the two studies which involved national surveys, services are reported to be variable and often limited. For neonatal units, this was reflected by the variability in both access to interpreting services and the availability of psychological support. (21) For hospital support palliative care teams, over two-thirds (64%) indicated they did not provide any form of bereavement support. (20)

The type and format of the bereavement support and counselling services was not always deemed to be needed or suitable for ethnic minority communities. One qualitative study reported that from the 18 participants, 17 found their family was their main source of support during their bereavement.(25) Friends, neighbours and support from their religious communities were also commonplace, with just four indicating that the community specialist palliative care team support had been helpful during this time.(25) A further study, comparing bereaved people from a black Caribbean and white ethnicity backgrounds, emphasised the importance of personal faith and the support provided via their religious leader for the black Caribbean participants.(24) It was also noteworthy that the format of other methods of support, such as a memorial service, traditionally had a strong Christian influence. (20) Whilst this might have been in keeping with the main cultural background from that specific local community, this may have precluded those from other faiths or cultures from attending. (20) Practical legal and financial support was often recognised to be needed rather than specialist interventions. One study indicated that socio-economic factors, such as financial worries, legal and housing problems, were reported more commonly by black Caribbean respondents compared with white respondents, during the bereavement period.(23) These factors were associated with a higher prevalence of anxiety and depression among black Caribbean respondents.(23) A further study also reported on the significant financial difficulties experienced by half of the 19 Bangladeshi participants after the death e.g. trying to meet the costs of transporting the deceased back to Bangladesh.(25)

An additional barrier to accessing bereavement care related to culturally specific belief
s. Within Romany Gypsy and Traveller culture, for example, there is a practice of ‘not speaking about’ bereavement within the close-knit community.(22) This internalisation can potentially increase the risk to poor mental health following significant loss or bereavement. There is reported stigma and shame associated with mental health illness, including bereavement-related depression, resulting in a reluctance to seek formal support.(22)

#### Facilitators

The overarching theme was ‘accessibility’ with the two subthemes being ‘readily available information’ and ‘inclusive approaches’. One study, in the context of support for bereaved parents, reported that information about the needs of different faith groups was easily available for healthcare professionals to access.(21) Additionally, 78% of participants reported that contact details for representatives of different faith groups were available.(21)

A further study, in the context of an invite to a memorial service, suggested that adopting an inclusive invite approach to support was important rather than selecting specific individuals (e.g. only those who had received input from the hospice team).(20)

#### Models of care provision

There were three studies which reported on examples of different models of care provision. One study reported on the role of a bilingual health-care worker in Bradford, based with the local community and hospital-based specialist palliative care teams, to help facilitate links with the South Asian community.(19) Although, the remit was wide, the healthcare worker had a key role in communication and family support prior to death and specifically supported one of the seventeen families during the bereavement period.(19) Their important role in terms of having a similar cultural background, and hence an appreciation and understanding of beliefs, practices and the expression of grief, was also acknowledged.(19)

A further study reported on the use of a memorial service for bereaved relatives of palliative care patients who had died within an acute hospital. The aim of this model was to try to address the needs of individuals and families who may not be offered any alternative means of bereavement support. (20) Within the third study, which focused on neonatal units, memorial or remembrance services were also a recognised model of support for bereaved parents.(21)

#### Outcomes from those who access bereavement services

There were no identified studies reporting outcomes from ethnic minority groups accessing bereavement support and counselling services. The study focused on the role of the bilingual health-care worker was a retrospective review and no direct views from services users were obtained.(19) It was also noteworthy that no formal evaluation of memorial services had been conducted and that planned evaluations tended to be in the format of audits.(20)

#### Levels of satisfaction with bereavement services

There were no identified studies reporting levels of satisfaction with bereavement care, support and counselling ethnic minority groups.

#### Conceptual map

An overview of the preliminary conceptual map derived from the study findings (*Figure 2*) outlines findings from included studies relating to the facilitators, barriers, outcomes and satisfaction relating to bereavement care for ethnic minority groups. This was constructed using the limited research literature available, but it conveys what is available to date.

**Figure 2:**
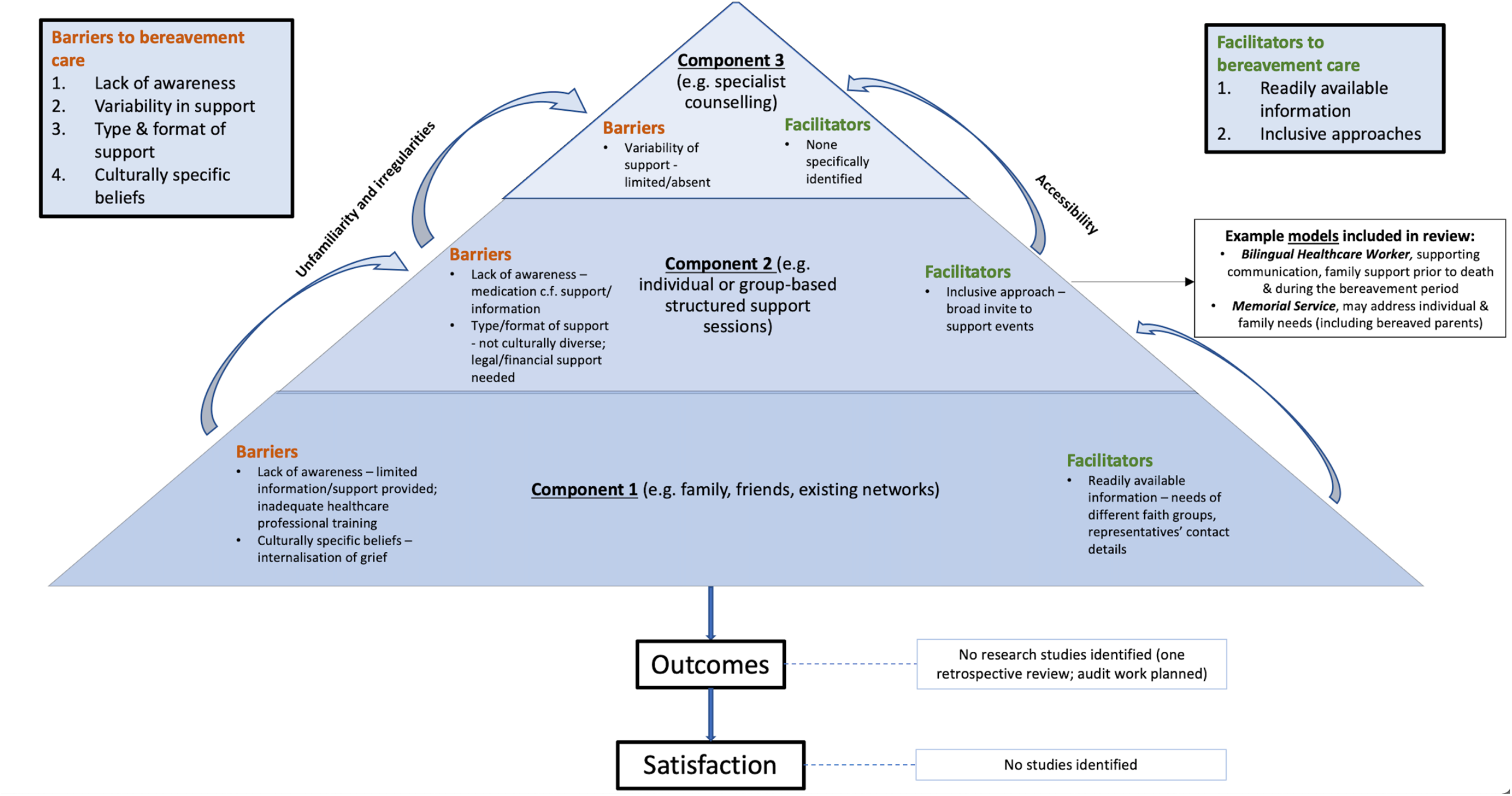
A preliminary conceptual map of existing facilitators, barriers, outcomes and satisfaction relating to bereavement care for ethnic minority groups based on findings from included studies.

## Discussion

### Summary of main findings

This review reveals a stark lack of evidence about bereavement care for ethnic minority populations. There is no research literature outlining the role of family, friends and existing networks, other than the suggestion that this type of support, including the role of religious communities and faith, is especially important. From the limited evidence available, there are barriers at each level of the three identified components of bereavement care outlined by the NBA for ethnic minority groups, limiting accessibility. In particular, issues relating to the availability, awareness and dissemination of information were identified, which ideally should be available on a universal basis; furthermore, barriers at components one and two may also impact on awareness and access to bereavement counselling. A lack of relevant, culturally competent training for healthcare professionals can limit access and awareness of potential support services. Additionally, these services may not be structured in a way which meets the needs of ethnic minority groups. For example, practical, legal and financial support may be needed and deemed more important by users during the bereavement period. There were few examples of existing models of care, a real absence of evidence about outcomes and levels of satisfaction for those from an ethnic minority background who receive bereavement care and no identified studies which focused on users who were children.

### Comparison with existing literature

The challenges seen in ensuring adequate access to bereavement care resonates with findings from both the recent ethnic minority mapping report(8) and a previous literature and evidence review focused on racial disparities for mental health services.(26) The latter report found those from black and minority ethnic communities were less likely to access mental health support through their General Practitioner and were more likely to end up in crisis care. Additionally, it is recognised there is a low uptake from ethnic minority groups of Specialist Palliative Care (SPC) services within the UK.(27) Potential reasons are complex and may include a lack of understanding about appropriate referral criteria, lack of awareness or understanding about services, conflicting ideas with the philosophy of hospice and palliative care, previous poor experience, and geographical barriers relating to the location of hospices.(27)(28)(29)

These findings have wider implications and potential impact on bereavement experience. It is recognised that the quality of care and support prior to death, has subsequent impact on those who are bereaved. For example, a national post-bereavement survey within Japan, reported that having good end-of-life care discussions was associated with a reduced likelihood of experiencing complicated grief.(30) If access to SPC services is limited for those who are experiencing more complex issues prior to death, these individuals may not receive adequate specialist support in a timely manner both before and after the bereavement. Although SPC services are generally seen as providers of bereavement counselling, they can offer a variety of bereavement care spanning all three components such as individual or group-based structured support sessions. By way of comparison, poor levels of satisfaction with SPC services can be associated with more bereavement related issues.(27)

Issues relating to the design and structure of services has also been described within a previous study focused on loneliness and isolation for ethnic minority groups.(31) Additionally, social isolation for different ethnic minority older people has also been explored.(32) Despite evidence of need, perceptions from bereaved ethnic minority individuals reflected care was not culturally sensitive or designed with their needs in mind e.g. a preference for informal ‘drop in’ support.(31)

### Implications for research and practice

Future research needs a much greater focus on ethnic minority communities to develop an evidence-base for the provision of bereavement care. In part, this means the recording of relevant data needs to be more comprehensive and complete.(33) Equally, if not more importantly, participatory or co-design methodological approaches are needed.(34) Collaborative studies with key stakeholders from the relevant communities are required to build meaningful alliances and facilitate social change, such as those which have been undertaken with the Roma community to try to overcome health inequalities.(35) Full engagement is needed to ensure culture and beliefs are reflected, respected and guide the development of appropriate bereavement care. Careful consideration as to whom may be key ‘door openers’ is required. This may include ethnic minority health advocates and faith leaders; building trusting relationships is required.

Key learning from the model of social prescribing,(36) which involves link workers providing non-medical support for health and well-being, has relevance in the context of bereavement care. Link workers could provide knowledge and help individuals navigate the different systems and mechanism of support. As well as connections to local community groups, this may also help connections with statutory services providing relevant legal, housing and financial information.

Aligning with the philosophy and modelling of ‘compassionate communities’ would be another example. Communities are encouraged to ‘support people and their families who are dying or living with loss’ and provides a linkage between professional healthcare and organic supportive networks within the community.(37) Communication is a key factor in enhancing quality of care. Within the National Ambitions for Palliative and End-of-life Care, the first priority is focused on individualised care and the ability to have honest conversations.(38) It is recognised, however, that those not being able to speak the dominant language e.g. English, can face challenges in accessing palliative care and issues relating to communication between patients, family carers and healthcare professionals can lead to dissatisfaction with care.(28) In the context of a public health approach to support bereavement care, upskilling and enabling relevant community workers (such as food bank employees, community support workers, hairdressers) (39) to be aware of, and feel competent to provide, component one bereavement support i.e. listening and signposting to additional support may be pertinent. This is not overlooking healthcare professionals, such as General Practitioners, who continue to have a key role to play in minimising barriers to accessing bereavement care. Efforts need to ensure that relevant information is available, accessible, and signposting facilitated to support services which are in keeping with cultural needs.

Further exploration, directly from users, about the most appropriate models and format of support needs to be obtained so that factors important to ethnic minority communities are incorporated into service design and delivery. This links with the recommendations from the recent report focused on the needs of ethnic minority communities arising from the COVID-19 pandemic, which include: i) establishing a national Black, Asian and minority ethnic (BAME) Bereavement Service that meets the cultural needs of the community; ii) bereavement therapists and service providers to have cultural competency training that is quality assured; and iii) the identification of good practice in addressing the disparity of bereavement and loss support for BAME communities using an intersectional approach. In turn, research needs to focus, not just on what is needed, but robust evaluations of the efficacy and satisfaction with different models of care for both adults and bereaved children from ethnic minorities communities.

### Strengths and limitations

We adopted a rigorous approach to extracting, searching and appraising the existing literature, working across a multi-disciplinary team and embedding engagement with patient and key stakeholder representatives. Furthermore, we developed a comprehensive search strategy (including terms such as ‘refugees’) and extracted articles from research databases and those containing grey literature (e.g. Trip database and ProQuest). Included studies comprised descriptive, quantitative non-randomised and qualitative studies. Whilst these study designs are typically aligned with lower levels of evidence,(40) we are confident in their findings given a broad appraisal of good quality across the included studies.

Our searching of grey literature, however, could have been more extensive if additional hand-searching had been undertaken (potentially using key authors names) and the inclusion of relevant research reported within books. Additionally, we didn’t include specific search terms focused on the concept of ‘a good death’ which may have had relevance about the impact on bereavement care. We focused on UK studies and the literature obtained tended to have a health-based focus. In view of these factors, some sources of data may have been overlooked.

## Conclusions

There is a stark lack of an evidence-base to support UK bereavement care for ethnic minority communities. Our initial, heuristic findings suggest there is a pressing need to understand the role in bereavement played by families, friends, individual religious beliefs and faith networks and other community support systems in aiding those from diverse ethnic minority backgrounds. This needs to be considered in the context of changing demographics and prevalence of specific disease. Fundamental to this agenda will be the effective and meaningful engagement of people from ethnic minority communities in the co-design and interpretation of future research, to produce findings that can inform and improve service assess, innovate models of service provision and support, and increase satisfaction with those services among the bereaved families.

## Data Availability

All data is included in the manuscript. Further information about the search strategies is available from the corresponding author on reasonable request.

## Acknowledgements

We wish to acknowledge the following individuals for their time and thoughtful contributions: Jane McCarthy; Alison Penny; Sabeen Zahra; Debjani Chatterjee and Marilyn Relf.

## Authorship

CRM, RAP and MJA conceived and designed the study. MJA, GC and BE completed all the searches and conducted the initial screening and full manuscript reviews. CRM and MJA analysed and interpreted the data. CRM and MJA drafted the manuscript and all authors have read and approved the submitted version.

## Data availability statement

Further information about the search strategies are available from the corresponding author on reasonable request.

---

**Supplementary file 1: Example of search strategy used in Medline**

Database: Ovid MEDLINE(R) and Epub Ahead of Print, In-Process & Other Non-Indexed Citations, Daily and Versions(R) <1946 to August 18, 2020>

Search Strategy:

--------------------------------------------------------------------------------

1. exp bereavement/ (13314)
2. (bereave* or grief or mourn*).mp. [mp=title, abstract, original title, name of substance word, subject heading word, floating sub-heading word, keyword heading word, organism supplementary concept word, protocol supplementary concept word, rare disease supplementary concept word, unique identifier, synonyms] (19259)
3. Attitude to Death.mp. or exp Attitude to Death/ (16173)
4. or/1-3 [bereavement] (32698)
5. exp Ethnic Groups/ (153675)
6. Minority Groups/ (13934)
7. Refugees/ (10245)
8. exp Continental Population Groups/ (220470)
9. (black? or ethnic* or minority or minorities or BAME).mp. [mp=title, abstract, original title, name of substance word, subject heading word, floating sub-heading word, keyword heading word, organism supplementary concept word, protocol supplementary concept word, rare disease supplementary concept word, unique identifier, synonyms] (353288)
10. (multi?cultural or multi cultural or cross?cultural or cross cultural or trans?cultural or transcultural).mp. [mp=title, abstract, original title, name of substance word, subject heading word, floating sub-heading word, keyword heading word, organism supplementary concept word, protocol supplementary concept word, rare disease supplementary concept word, unique identifier, synonyms] (40152)
11. exp “Emigrants and Immigrants”/ (12483)
12. (emigrant* or immigrant* or migrant* or refugee*).mp. [mp=title, abstract, original title, name of substance word, subject heading word, floating sub-heading word, keyword heading word, organism supplementary concept word, protocol supplementary concept word, rare disease supplementary concept word, unique identifier, synonyms] (61370)
13. or/5-12 [BAME] (601049)
14. (islam* or muslim* or hindu* or sikh* or buddhism* or jew*).mp. [mp=title, abstract, original title, name of substance word, subject heading word, floating sub-heading word, keyword heading word, organism supplementary concept word, protocol supplementary concept word, rare disease supplementary concept word, unique identifier, synonyms] (30776)
15. buddhism/ or hinduism/ or islam/ or judaism/ (9348)
16. 5 or 6 or 7 or 8 or 9 or 10 or 11 or 12 or 14 or 15 [BAME & religion] (620463)
17. exp United Kingdom/ (365573)
18. (national health service* or nhs*).mp. [mp=title, abstract, original title, name of substance word, subject heading word, floating sub-heading word, keyword heading word, organism supplementary concept word, protocol supplementary concept word, rare disease supplementary concept word, unique identifier, synonyms] (44331)
19. (english not ((published or publication* or translat* or written or language* or speak* or literature or citation*) adj5 english)).mp. [mp=title, abstract, original title, name of substance word, subject heading word, floating sub-heading word, keyword heading word, organism supplementary concept word, protocol supplementary concept word, rare disease supplementary concept word, unique identifier, synonyms] (1597583)
20. (gb or “g.b.” or britain* or (british* not “british columbia”) or uk or “u.k.” or united kingdom* or (england* not “new england”) or northern ireland* or northern irish* or scotland* or scottish* or ((wales or “south wales”) not “new south wales”) or welsh*).mp. [mp=title, abstract, original title, name of substance word, subject heading word, floating sub-heading word, keyword heading word, organism supplementary concept word, protocol supplementary concept word, rare disease supplementary concept word, unique identifier, synonyms] (488332)
21. (bath or “bath’s” or ((birmingham not alabama*) or (“birmingham’s” not alabama*) or bradford or “bradford’s” or brighton or “brighton’s” or bristol or “bristol’s” or carlisle* or “carlisle’s” or (cambridge not (massachusetts* or boston* or harvard*)) or (“cambridge’s” not (massachusetts* or boston* or harvard*)) or (canterbury not zealand*) or (“canterbury’s” not zealand*) or chelmsford or “chelmsford’s” or chester or “chester’s” or chichester or “chichester’s” or coventry or “coventry’s” or derby or “derby’s” or (durham not (carolina* or nc)) or (“durham’s” not (carolina* or nc)) or ely or “ely’s” or exeter or “exeter’s” or gloucester or “gloucester’s” or hereford or “hereford’s” or hull or “hull’s” or lancaster or “lancaster’s” or leeds* or leicester or “leicester’s” or (lincoln not nebraska*) or (“lincoln’s” not nebraska*) or (liverpool not (new south wales* or nsw)) or (“liverpool’s” not (new south wales* or nsw)) or ((london not (ontario* or ont or toronto*)) or (“london’s” not (ontario* or ont or toronto*)) or manchester or “manchester’s” or (newcastle not (new south wales* or nsw)) or (“newcastle’s” not (new south wales* or nsw)) or norwich or “norwich’s” or nottingham or “nottingham’s” or oxford or “oxford’s” or peterborough or “peterborough’s” or plymouth or “plymouth’s” or portsmouth or “portsmouth’s” or preston or “preston’s” or ripon or “ripon’s” or salford or “salford’s” or salisbury or “salisbury’s” or sheffield or “sheffield’s” or southampton or “southampton’s” or st albans or stoke or “stoke’s” or sunderland or “sunderland’s” or truro or “truro’s” or wakefield or “wakefield’s” or wells or westminster or “westminster’s” or winchester or “winchester’s” or wolverhampton or “wolverhampton’s” or (worcester not (massachusetts* or boston* or harvard*)) or (“worcester’s” not (massachusetts* or boston* or harvard*)) or (york not (“new york*” or ny or ontario* or ont or toronto*)) or (“york’s” not (“new york*” or ny or ontario* or ont or toronto*))))).mp. [mp=title, abstract, original title, name of substance word, subject heading word, floating sub-heading word, keyword heading word, organism supplementary concept word, protocol supplementary concept word, rare disease supplementary concept word, unique identifier, synonyms] (198503)
22. (bangor or “bangor’s” or cardiff or “cardiff’s” or newport or “newport’s” or st asaph or “st asaph’s” or st davids or swansea or “swansea’s”).mp. [mp=title, abstract, original title, name of substance word, subject heading word, floating sub-heading word, keyword heading word, organism supplementary concept word, protocol supplementary concept word, rare disease supplementary concept word, unique identifier, synonyms] (3042)
23. (aberdeen or “aberdeen’s” or dundee or “dundee’s” or edinburgh or “edinburgh’s” or glasgow or “glasgow’s” or inverness or (perth not australia*) or (“perth’s” not australia*) or stirling or “stirling’s”).mp. [mp=title, abstract, original title, name of substance word, subject heading word, floating sub-heading word, keyword heading word, organism supplementary concept word, protocol supplementary concept word, rare disease supplementary concept word, unique identifier, synonyms] (39571)
24. (armagh or “armagh’s” or belfast or “belfast’s” or lisburn or “lisburn’s” or londonderry or “londonderry’s” or derry or “derry’s” or newry or “newry’s”).mp. [mp=title, abstract, original title, name of substance word, subject heading word, floating sub-heading word, keyword heading word, organism supplementary concept word, protocol supplementary concept word, rare disease supplementary concept word, unique identifier, synonyms] (1460)
25. or/17-24 [UK context] (2276840)
26. (exp africa/ or exp americas/ or exp antarctic regions/ or exp arctic regions/ or exp asia/ or exp australia/ or exp oceania/) not (exp great britain/ or europe/) (2881116)
27. (“VOICES” or “Bereavement UK” or “National Survey of Bereaved People” or “child bereavement UK” or “National Bereavement Alliance” or “Cruse” or “Cruse Bereavement Care” or “Age UK” or “British Red” or “Dying Matters” or “Bereavement Services Association”).mp. [mp=title, abstract, original title, name of substance word, subject heading word, floating sub-heading word, keyword heading word, organism supplementary concept word, protocol supplementary concept word, rare disease supplementary concept word, unique identifier, synonyms] (8018)
28. 25 and 26 (148226)
29. 25 or 27 (2283892)
30. 4 and 16 and 29 (251)

**Supplementary file 2:**
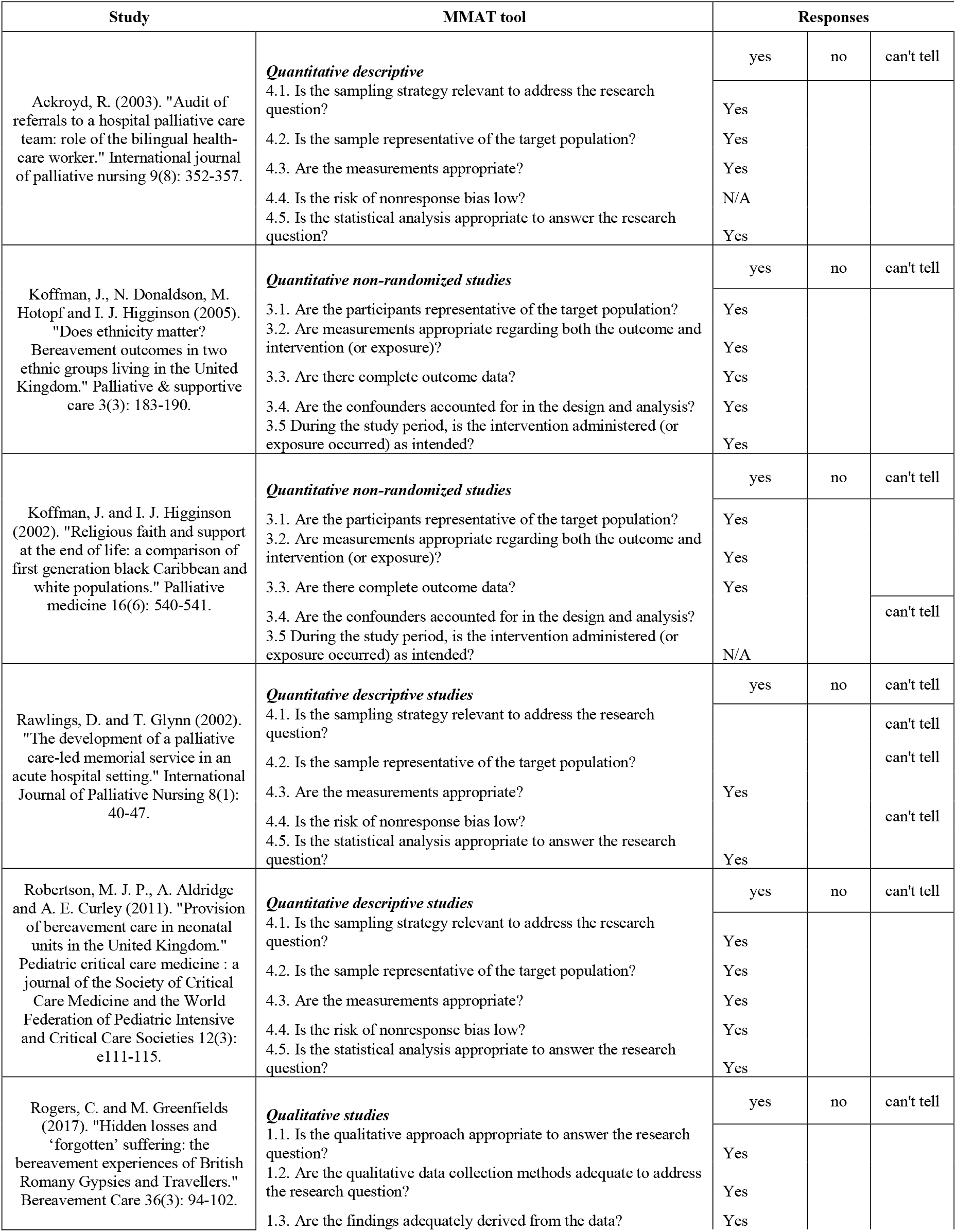
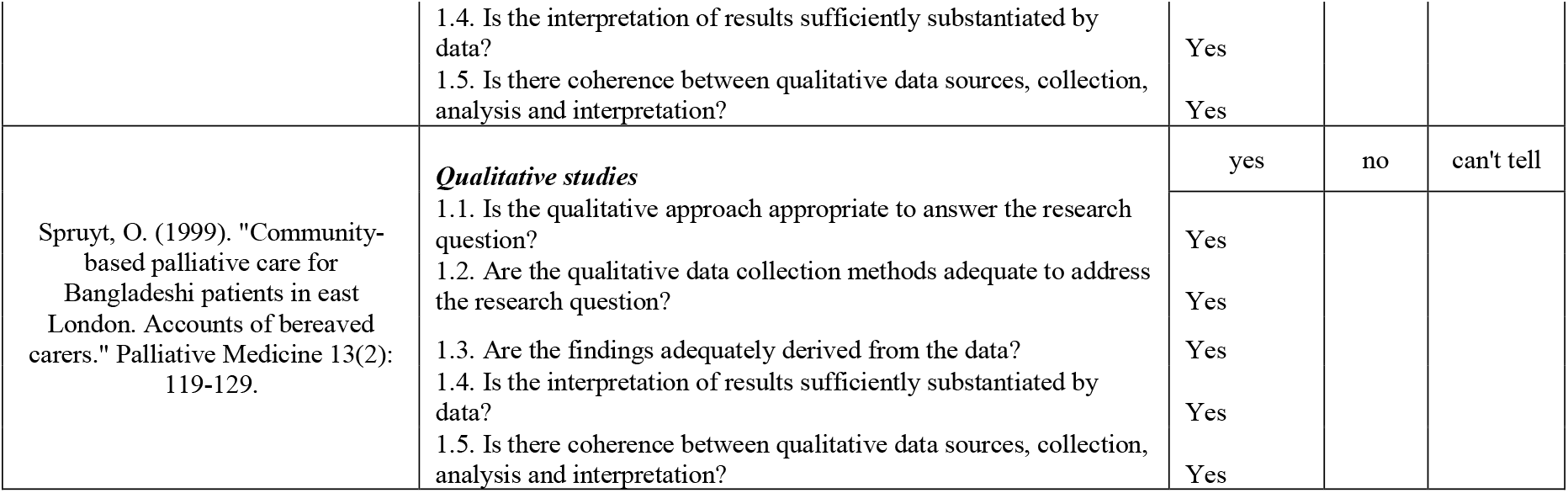
MMAT quality appraisal summary.

## References

1. COVID-19 Daily Deaths [Internet]. 2021. Available from: https://www.england.nhs.uk/statistics/wp-content/uploads/sites/2/2021/01/COVID-19-daily-announced-deaths-2-January-2021.xlsx.

2. Elwell-Sutton T, Deeny S, Stafford M. Emerging findings on the impact of COVID-19 on black and minority ethnic people. https://www.health.org.uk/news-and-comment/charts-and-infographics/emerging-findings-on-the-impact-of-covid-19-on-black-and-min: The Health Foundation; 2020.

3. Coronavirus (COVID-19) related deaths by ethnic group, England and Wales: 2 March 2020 to 15 May 2020 [Internet]. 2020 [cited 1st October 2020]. Available from: https://www.ons.gov.uk/peoplepopulationandcommunity/birthsdeathsandmarriages/deaths/articles/coronaviruscovid19relateddeathsbyethnicgroupenglandandwales/2march2020to15may2020.

4. Singer M, Bulled N, Ostrach B, Mendenhall E. Syndemics and the biosocial conception of health. The Lancet. 2017;389(10072):941–50.

5. Stroebe M, Schut H. Bereavement in Times of COVID-19: A Review and Theoretical Framework. OMEGA - Journal of Death and Dying. 2020;82(3):500–22.

6. Selman LE, Chao D, Sowden R, Marshall S, Chamberlain C, Koffman J. Bereavement Support on the Frontline of COVID-19: Recommendations for Hospital Clinicians. Journal of Pain and Symptom Management. 2020;60(2):e81–e6.

7. Mayland CR, Harding AJE, Preston N, Payne S. Supporting Adults Bereaved Through COVID-19: A Rapid Review of the Impact of Previous Pandemics on Grief and Bereavement. Journal of Pain and Symptom Management. 2020;60(2):e33–e9.

8. Murray K. National Mapping of BAME Mental Health Services. London: BAMEstream; 2020.

9. Cross BR. Barriers to belonging. An exploration of loneliness among people from Black, Asian and Minority Ethnic backgrounds.

10. Rumbold B, and Aoun, Samar. Bereavement and palliative care: A public health perspective. Progress in Palliative Care 2014;22(3):132–5.

11. O’Connor MF, Arizmendi BJ. Neuropsychological correlates of complicated grief in older spousally bereaved adults. J Gerontol B Psychol Sci Soc Sci. 2014;69(1):12–8.

12. Penny A, Relf M. A Guide to Commissioning Bereavement Services in England. London: National Bereavement Alliance 2017.

13. Aoun SM, Breen LJ, Rumbold B, Howting D. Reported experiences of bereavement support in Western Australia: a pilot study. Australian and New Zealand Journal of Public Health. 2014;38(5):473–9.

14. Evans R, Ribbens McCarthy J, Kébé F, Bowlby S, Wouango J. Interpreting ‘grief’ in Senegal: language, emotions and cross-cultural translation in a francophone African context. Mortality. 2017;22(2):118–35.

15. Brunton G, Oliver S, Thomas J. Innovations in framework synthesis as a systematic review method. Research Synthesis Methods. 2020;11(3):316–30.

16. Moher D, Liberati A, Tetzlaff J, Altman DG, The PG. Preferred Reporting Items for Systematic Reviews and Meta-Analyses: The PRISMA Statement. PLOS Medicine. 2009;6(7):e1000097.

17. Hong QN, Fàbregues S, Bartlett G, Boardman F, Cargo M, Dagenais P, et al. The Mixed Methods Appraisal Tool (MMAT) version 2018 for information professionals and researchers. Education for Information. 2018;34:1–7.

18. Dixon J, King D, Matosevic T, Clark M, Knapp M. Equity in the provision of palliative care in the UK: review of evidence. Marie Curie/ PSSRU, LSE2015.

19. Ackroyd R. Audit of referrals to a hospital palliative care team: role of the bilingual health-care worker. Int J Palliat Nurs. 2003;9(8):352–7.

20. Rawlings D, Glynn T. The development of a palliative care-led memorial service in an acute hospital setting. International Journal of Palliative Nursing. 2002;8(1):40–7.

21. Robertson MJ, Aldridge A, Curley AE. Provision of bereavement care in neonatal units in the United Kingdom. Pediatr Crit Care Med. 2011;12(3):e111–5.

22. Rogers C, Greenfields M. Hidden losses and ‘forgotten’ suffering: the bereavement experiences of British Romany Gypsies and Travellers. Bereavement Care. 2017;36(3):94–102.

23. Koffman J, Donaldson N, Hotopf M, Higginson IJ. Does ethnicity matter? Bereavement outcomes in two ethnic groups living in the United Kingdom. Palliative and Supportive Care. 2005;3(3):183–90.

24. Koffman J, Higginson IJ. Religious faith and support at the end of life: a comparison of first generation black Caribbean and white populations. Palliative Medicine. 2002;16(6):540–1.

25. Spruyt O. Community-based palliative care for Bangladeshi patients in east London. Accounts of bereaved carers. Palliat Med. 1999;13(2):119–29.

26. Bignall T, Jeraj S, Helsby E, Butt J. Racial disparities in mental health: Literature and evidence review. London: Race Equality Foundation 2019.

27. Calanzani N, Koffman J, Higginson IJ. Palliative and end of life care for Black, Asian and Minority Ethnic groups in the UK. London: Marie Curie Cancer Care 2013.

28. Gunaratnam Y. Improving the quality of palliative care. London: Race Equality Foundation; 2007.

29. Philips L, Taylor V. Addressing the palliative care needs of minority groups. Primary Health Care. 2012;22(1):26–30.

30. Yamaguchi T, Maeda I, Hatano Y, Mori M, Shima Y, Tsuneto S, et al. Effects of End-of-Life Discussions on the Mental Health of Bereaved Family Members and Quality of Patient Death and Care. J Pain Symptom Manage. 2017;54(1):17-26.e1.

31. British Red Cross Society. Barriers to belonging: An exploration of loneliness among people from Black, Asian and Minority Ethnic backgrounds. London: British Red Cross Society; 2019.

32. Yarker S. Ageing in Place for Minority Ethnic ommunities: The importance of social infrastructure 2020.

33. Zhang X, Pérez-Stable EJ, Bourne PE, Peprah E, Duru OK, Breen N, et al. Big Data Science: Opportunities and Challenges to Address Minority Health and Health Disparities in the 21st Century. Ethn Dis. 2017;27(2):95–106.

34. Balezdrova N, Choi D, Lam B. ‘Invisible Minorities’: Exploring Improvement Strategies for Social Care Services aimed at Elderly Immigrants in the UK using Co-Design Methods 2019.

35. Miranda DE, García-Ramírez M, Albar-Marín MJ. Building Meaningful Community Advocacy for Ethnic-based Health Equity: The RoAd4Health Experience. American Journal of Community Psychology. 2020;n/a(n/a).

36. South J, Higgins TJ, Woodall J, White SM. Can social prescribing provide the missing link? Primary Health Care Research & Development. 2008;9(4):310–8.

37. Barry V, Patel M. An Overview of Compassionate Communities in England. West Midlands, London: Murray Hall Community Trust, National Council for Palliative Care Dying Matters Coalition; 2013.

38. National Palliative and End of Life Care Partnership. Ambitions for Palliative and End of Life Care: A national framework for local action 2015-2020. London: National Palliative and End of Life Care Partnership; 2015.

39. ehospice. Hospice teams up with hair salon for bereavement awareness https://ehospice.com/uk_posts/hospice-teams-up-with-hair-salon-for-bereavement-awareness/: ehospice; 2019 [Available from: https://ehospice.com/uk_posts/hospice-teams-up-with-hair-salon-for-bereavement-awareness/.

40. The Oxford 2011 levels of evidence. [Internet]. 2021 [cited 4th January 2021]. Available from: http://www.cebm.net/index.aspx?o=5653

